# Precision Medicine in Education: Using Clustering Analysis for Personalized Student Support in Medical Training

**DOI:** 10.1101/2025.09.09.25335466

**Authors:** Ardo Sanjaya, Christian Edwin, Cindra Paskaria, Nathanael Andry Mianto, Renata Alviani, Kevin Gunawan

## Abstract

**Background:** Burnout, stress, and psychological distress are prevalent among medical students, undermining academic performance and well-being. Traditional one-size-fits-all interventions have shown limited effectiveness, highlighting the need for precision student support. This study applied unsupervised learning to classify medical students into clusters and examined their academic and psychological characteristics.

**Methods:** A cross-sectional study was conducted among 677 medical students. Psychological distress (DASS-21), burnout (SBI), resilience (ARS-24), coping strategies (Brief-COPE), and learning environment perception (DREEM-12) were assessed. Spectral clustering identified four profiles, validated through resampling. ANCOVA compared GPA and psychological measures across clusters, adjusting for covariates. Focus group discussions provided qualitative validation.

**Results:** Four clusters were identified. Cluster C was the most at-risk, showing high distress, low resilience, and poor academic performance. Cluster A also reported high stress but buffered through resilience and adaptive coping. Cluster B exhibited the strongest resilience and lowest stress. Cluster D achieved the highest GPA, but with moderate resilience and coping scores. ANCOVA confirmed significant differences (p < 0.001) across clusters, independent of covariates. Qualitative findings reinforced these profiles, highlighting mechanisms such as stigma reframed as motivation (Cluster A), autonomous and thriving (Cluster B), lack of autonomy and maladaptive coping (Cluster C), and fairness-sensitive students (Cluster D).

**Conclusion:** Clustering analysis reveals psychologically distinct student groups with distinct support needs. Across clusters, precision student support should cultivate autonomy, resources, fairness, and scaffolding, aligning interventions with subgroup characteristics. These insights can inform tailored strategies to improve academic performance, reduce burnout, and foster resilience in medical education.

## Introduction

The sustainability of the medical workforce is a global concern that has gained increased attention recently. The World Health Organization emphasizes that supporting a resilient workforce of future healthcare professionals is crucial for current health demands and future challenges. A fundamental component of achieving this goal is the mental health of medical students. These students are expected to navigate the demanding requirements of studying medicine while experiencing high levels of burnout, stress, and exhaustion during their training. ^1–3^ Previous studies have shown that the increased burden during training may hamper academic performance and cause career dissatisfaction, possibly lowering their future performance. ^2,4,5^ Furthermore, the strain experienced during medical school also extends to medical practice, with medicine among the professions with the highest prevalence of burnout, depression, and chronic stress.^6–8^ These challenges may cause cascading effects extending beyond individual well-being, possibly affecting patient care and the sustainability of the healthcare systems. Left unaddressed, the mental health challenges could lead to a reduction in the number of individuals entering healthcare or even prompt those within healthcare to leave entirely. This battle of attrition diminished the available workforce and threatened effective healthcare services. Effective and targeted interventions are crucial to mitigate these risks.

Medical education, characterized by rigorous academic demands, unpredictable clinical scenarios, and considerable interpersonal stressors, is notoriously associated with a high prevalence of burnout and psychological distress.^9–12^ These challenges affect students’ emotional well-being and impair their learning, decision-making, and capacity for empathy in future practice. ^2^ Persistent stressors during education may also contribute to attrition or chronic psychiatric conditions such as depression and anxiety, compromising the future healthcare workforce.^7,13,14^ Therefore, identifying and supporting these students is essential not only for their benefits but also to sustain a competent healthcare workforce.

Traditional one-size-fits-all approaches to student support have shown, at best, moderate effects. ^15^ Studies have shown that targeted interventions demonstrated better overall reduction measures of psychological distress, such as stress. ^16^ However, due to the complex interplay of factors contributing to psychological distress in an educational environment, we may not be able to accurately identify students who may benefit from such interventions. Data-driven approaches utilizing unsupervised learning techniques offer a promising alternative by uncovering unseen patterns within complex datasets, supporting our understanding and classification of the complex experiences of medical students.

In particular, implementing unsupervised learning, clustering algorithms offer a promising approach to dissecting the complex nature of students’ psychological profiles. Since this approach did not require predefined labels, this technique can uncover patterns in data without prior assumptions, suitable for exploratory analysis. Several recent studies in educational settings have utilized clustering analysis to classify students based on their academic performance and engagement ^17–20^, or through large databases such as PISA scores. ^21^ However, the application of clustering analysis on mental health and psychological constructs in the context of academic performance remains underexplored.

Combining cluster analysis with analyzing students’ psychological and environmental data can help detect high-risk student clusters early. Utilizing several variables that affect academic performance, such as burnout ^22,23^, stress, learning environment perceptions ^1,24,25^, and coping ^26^ may reveal distinct student populations within the larger student body. This segmentation is important in designing and implementing targeted interventions that are effective and scalable. For instance, while personalized psychotherapy may be too expensive to apply across the entire student population, it could serve as a highly effective intervention for specific at-risk groups identified through clustering analysis. Recent clustering analysis studies have identified distinct student profiles that may present actionable, targetable interventions that improve student well-being and academic outcomes.^17,19,20^

This article applies clustering analysis to identify risk profiles based on mental health and psychological constructs among medical students. By combining unsupervised learning with qualitative validation, we introduce the concept of precision student support, modeled after precision medicine, where interventions are tailored to distinct psychological and academic profiles. This approach moves beyond one-size-fits-all wellness programs toward data-driven, targeted strategies that address the diverse needs of students. In the context of rising global concerns about burnout and depression in medical trainees, precision student support offers a scalable and resource-efficient framework to strengthen resilience, sustain academic performance, and ensure the long-term sustainability of the healthcare workforce.

## Methods

This study is a cross-sectional, single-center study involving medical students across all academic years (Semesters 1, 3, 5, and 7). The target population comprised 1,012 students. The survey was conducted using the SurveyMonkey survey platform and administered onsite through their mobile phones or laptops. The survey received responses from 862 students (85.2% response rate); out of those who responded, 726 students completed the study. To minimize confounding effects, students who had retaken the semester were excluded from the final analysis. Therefore, we retain 677 students for the final statistical analysis. Participation in this survey was voluntary, and the students’ identities were anonymized.

### Data Collection

Student data were collected using several standardized instruments (Table 1). The instruments were picked due to their availability and validity after translation into the Indonesian Language. Additionally, demographic and lifestyle data that might influence the results were collected, including age, gender, average sleep time, average study time, average social media use, participation in student organizations or extracurricular activities, chronic disease, mental health diagnosis, alcohol use, and smoking behavior.

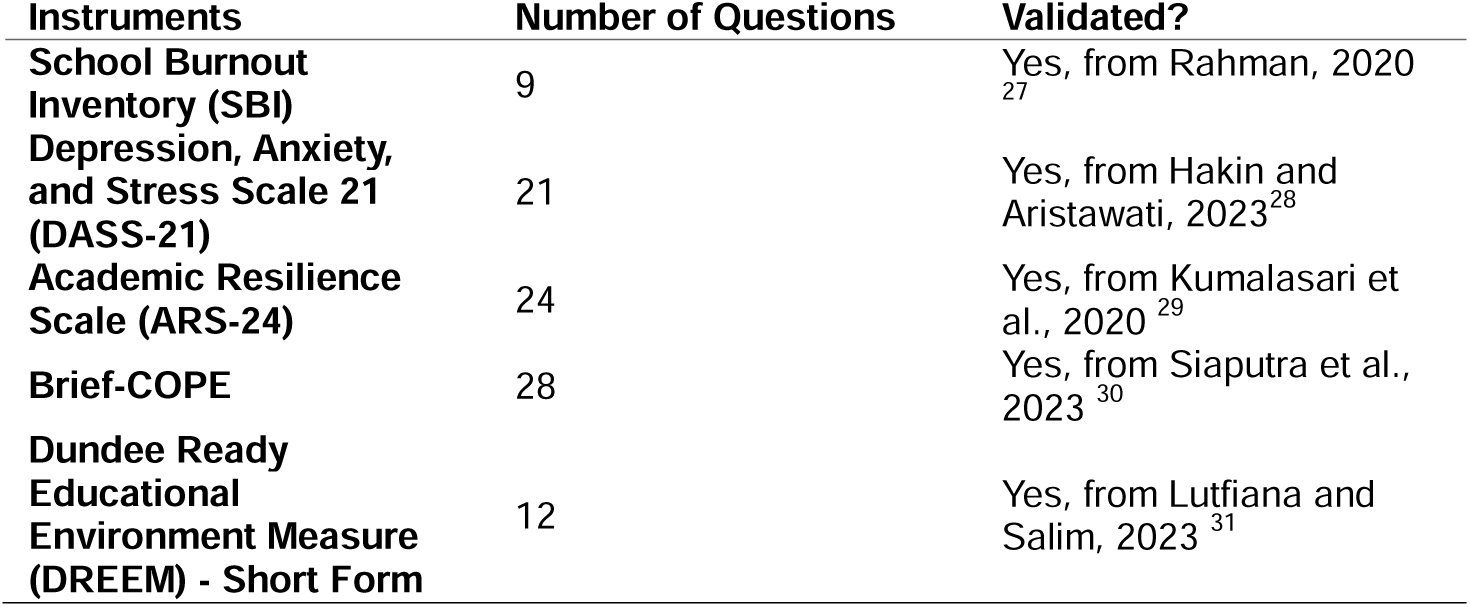

**Table 1.**
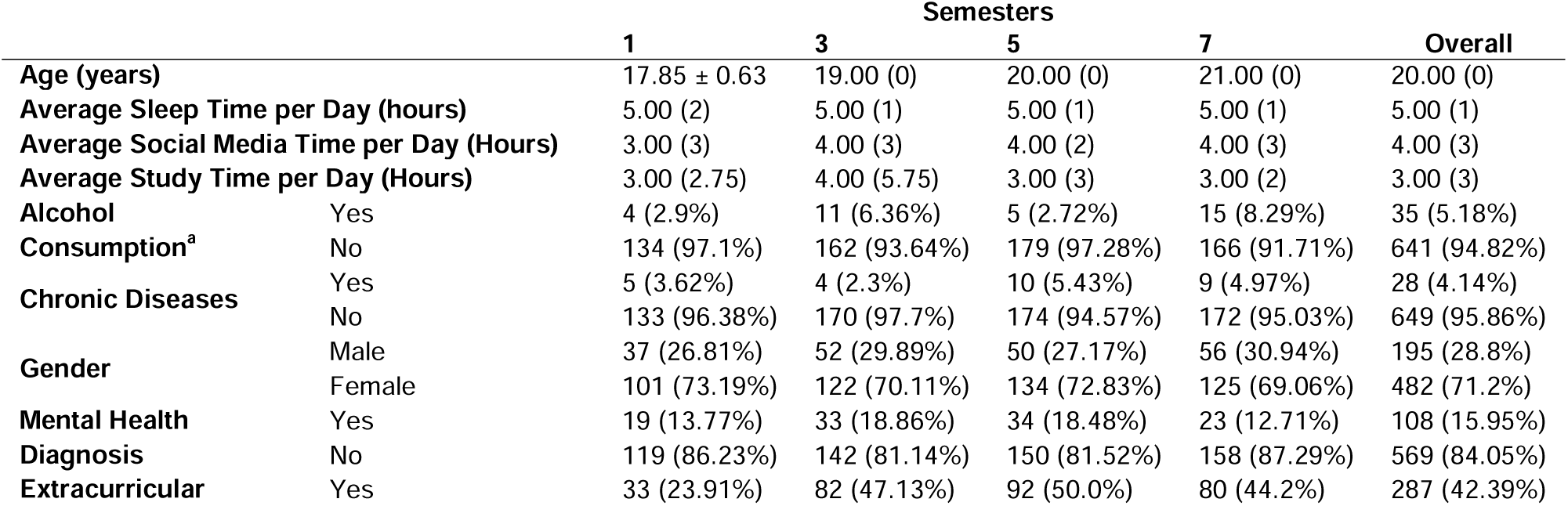
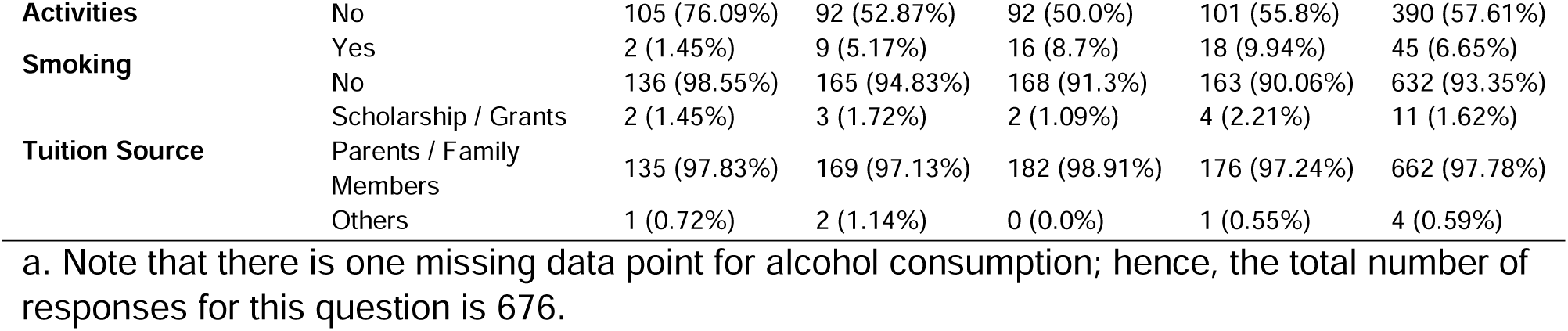
Subject Characteristics Across Semesters.

### Sample Demographics

Descriptive statistics were calculated and presented to summarize the demographic and contextual characteristics of the participants across the four groups. Variables assessed include age, average daily sleep time, social media usage, study time, and categorical variables such as alcohol consumption, chronic diseases, gender, mental health diagnosis, participation in extracurricular activities, smoking behavior, and tuition sources. Continuous variables were summarized using medians and interquartile ranges (IQR) or means and standard deviations based on normality testing, while categorical variables were presented as frequencies and percentages. Normality testing was done using the Shapiro-Wilk test, with p > 0.05 indicating normal distribution. These variables were evaluated as potential covariates in subsequent analysis.

### Clustering and Statistical Analysis

The clustering method used in this study is the Spectral Clustering method implemented through the Python package scikit-learn ^32^ (version 1.5.2). Data were limited to students from Semesters 3, 7, and 9 due to the unavailability of GPA scores for Semester 1 students (Remaining N = 540). We chose spectral clustering due to its ability to capture non-linear relationships, which should be better suited to capture the complexity of the psychological features in this dataset. Features were normalized by dividing their raw scores by the number of questions in each instrument or dimension to ensure comparability. Then, they were scaled to unit variance for the clustering algorithm. The optimal number of clusters was determined using the Davies-Bouldin Index (DBI) and Silhouette Score. We evaluated the cluster stability using a resampling-based approach with 1000 iterations, reporting the mean Adjusted Rand Index (ARI). The distinguishing features of each cluster were visualized using a heatmap of median values, and cluster separation was visualized using Principal Component Analysis (PCA). To ascertain that the clusters were distinct, an analysis of covariance (ANCOVA) was conducted to compare the features while controlling for confounding variables.

Assumptions of residual normality (Kolmogorov-Smirnov test), homogeneity of variances (Levene’s test), and homogeneity of regression slopes (interaction testing) were assessed before ANCOVA. ANCOVA was still conducted for minor deviations from normality. However, since some of the ANCOVA models were heteroscedastic, we implemented ANCOVA models with robust standard errors to account for heteroscedasticity. The independent variables were the Cluster information, while the dependent variables were the features used for clustering: SBI, DASS-21, and DREEM scores, along with subscales from the Brief-COPE (Problem-Focused, Emotion-Focused, and Avoidant Coping) and ARS-24 (Perseverance, Reflective and Adaptive Help-Seeking, and Negative Affect and Emotional Response). Academic performance, measured using grade point average (GPA), was also compared using ANCOVA. The covariates in our study include age, gender, average sleep time, average study time, average social media use, participation in student organizations or extracurricular activities, chronic disease, mental health diagnosis, alcohol use, and smoking behavior. We initially conducted a bivariate analysis of the covariate variables against each dependent variable. The final ANCOVA models included significant covariates from preliminary bivariate analyses to adjust for their influence. Significant covariates and their p-values and partial eta-squared values were calculated and presented in tabular format. Pairwise comparisons were conducted on significant ANCOVA results using Holm’s correction. All data visualization and statistical analyses were conducted using Python packages seaborn ^33^ (version 0.13.2) and statsmodels ^34^ (version 0.14.4).

### Qualitative Validation

To validate the clustering analysis, we conducted focus group discussions (FGDs) to explore the distinct psychological characteristics of the identified clusters. Two representative students were purposively selected from each academic year, for a total of eight students in each cluster. This approach ensures that all four clusters are represented across different stages of medical training. The qualitative sample was small and not intended to be representative of the entire cohort. The major aim of the qualitative component was to triangulate and contextualize the quantitative findings. Guided interviews were facilitated by two authors using a semi-structured protocol. The discussions were audio-recorded and transcribed verbatim using Plaud Note AI (Plaud AI, https://www.plaud.ai/; version 0095). Transcripts were re-checked manually for accuracy and validity. Several excerpts were translated into English and presented in the results section. Thematic analysis was carried out following Braun and Clarke’s six-phase framework. Two authors independently coded the transcripts and iteratively developed themes that captured the shared and divergent experiences of students across clusters. The qualitative findings were triangulated with the quantitative results to strengthen the validity of the cluster interpretations.

### Ethical Approval

This study adhered to the principles of the Declaration of Helsinki and received ethical approval from the Research Ethics Committee of Maranatha Christian University (Approval Number: 089/KEP/VII/2024). Electronic informed consent was obtained from all participants prior to their inclusion, with the consent form presented at the start of the online survey. Participants’ registration numbers, names, and email addresses were initially collected to facilitate accurate data matching and communication; however, all identifiable information was removed, and the dataset was fully anonymized before analysis to ensure confidentiality. Participants were informed that their responses would remain confidential and would be used exclusively for research purposes. For students who participated in focus group discussions, additional written informed consent was obtained. These participants agreed to have their interview data recorded, analyzed, and used anonymously in publications. All procedures for data collection and analysis complied with institutional and national ethical standards to safeguard participant privacy and well-being.

## Results

A total of 677 students, representing a 67% completion rate, were included in the final analysis. Demographic data of all the students in the analysis are shown in Table 1. Note that there are several variations of characteristics across semesters, such as the average study time per day, mental health diagnosis, and alcohol consumption, to name a few. The subjects consist primarily of female students (71.24%), and a significant portion (16.08%) have been diagnosed with mental health issues. Alcohol consumption, chronic disease diagnosis, and smoking were low in our subjects but varied with their current semester.

We conducted a clustering analysis to identify distinct psychological subgroups within the student population. Spectral Clustering was performed, and four clusters were selected (Figure 1A) based on a balance between the optimal values identified by Silhouette scores (3 clusters) and the Davies-Bouldin Index (5 clusters). We performed resampling with random subsets of 90% of the data to assess the stability of the clustering results, yielding a mean Adjusted Rand Index (ARI) of 0.68, indicating moderate stability. The resulting clusters (A–D) exhibited distinct psychological profiles and corresponding academic performance differences (Figure 1A). Cluster D comprised the highest-achieving students, as indicated by the highest GPA distribution, whereas Cluster C contained the lowest-achieving students (Figure 1B).

**Figure 1.**
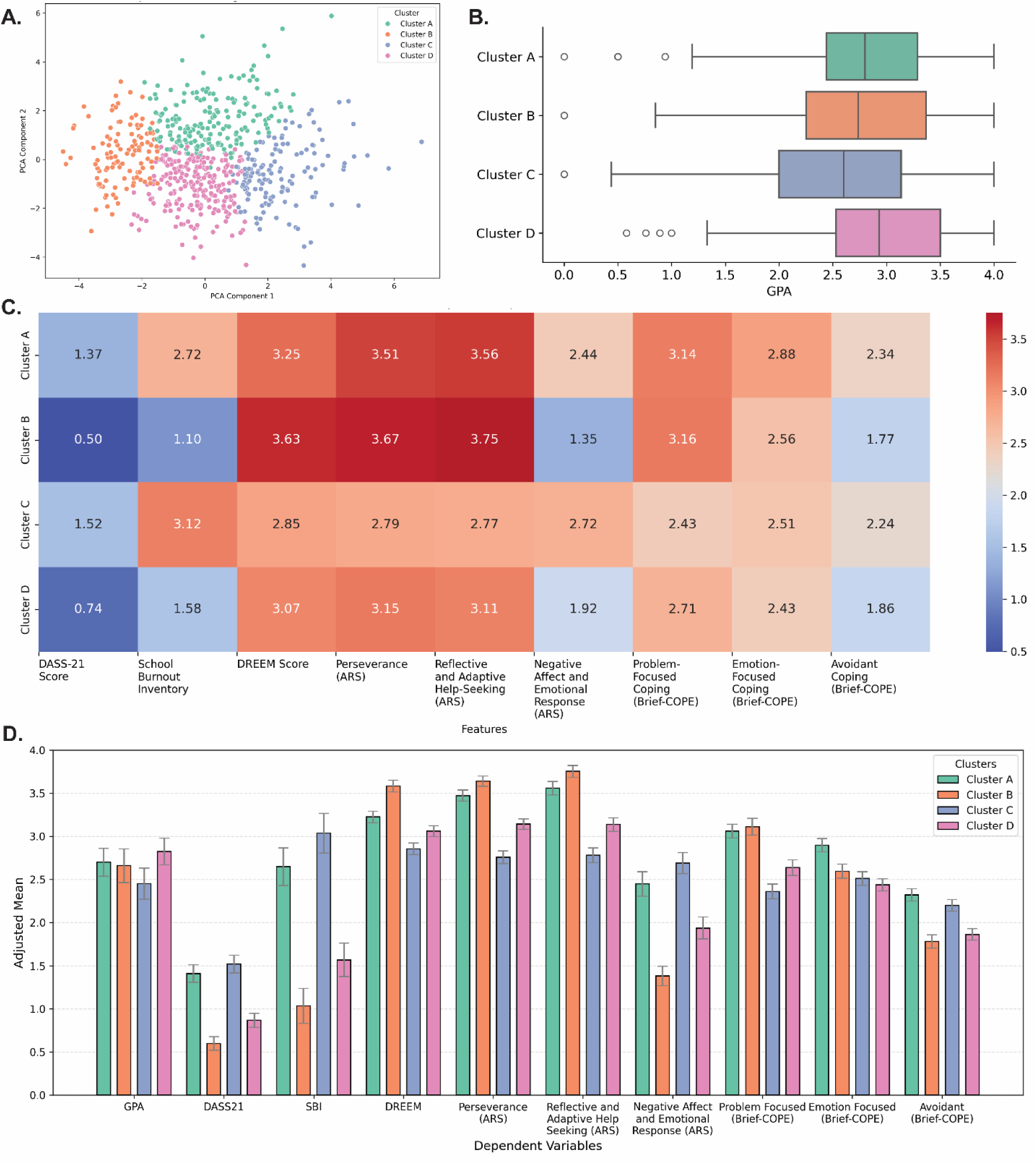
Clustering of Medical Students Based on Psychological Profiles and Learning Environment Perception. A. Scatter plot demonstrating the distribution of student clusters using principal component analysis. Each point represents a student being classified into four clusters (Cluster A, B, C, and D). B. Boxplot illustrating the distribution of GPA within each cluster. Cluster D has the highest median GPA value compared to the other clusters. Suggest cluster-specific differences in academic performance among the students. C. Heatmap showing the average scores of various psychological features, including depression, anxiety, stress, burnout, learning perception, academic resilience, and coping-related variables. Red colors indicate higher scores. Notice the pattern with clusters A and C as the clusters with the highest burnout and psychological stress, while clusters B and D are the clusters with the lowest burnout. Cluster B is unique as it represents students who feel highly supported and motivated to learn medicine. D. The bar plot shows the adjusted mean scores from ANCOVA models for various features across the four identified student clusters. Note that the clusters represent distinct feature differences even after controlling confounders, reinforcing the need for targeted interventions. Error bars denote the 95% confidence intervals for the adjusted mean.

Two main patterns emerged. Clusters A and C both showed high stress and burnout but diverged in resilience and coping. Cluster A maintained higher resilience and coping, while C had the weakest scores and perceived their learning environment most negatively. Clusters B and D reported lower stress overall, though B thrived with the strongest resilience and learning perceptions, whereas D remained moderate across domains. These contrasts highlight heterogeneity beyond what raw stress levels alone reveal (Figure 1C).

Our analysis revealed significant psychological and academic differences between student clusters, even after adjusting for covariates (p < 0.001 for all variables; Table 2). These differences cannot be explained solely by mental health history, sleep, extracurricular activities, or alcohol use, but instead reflect inherent psychological profiles. GPA differed significantly between clusters (F = 6.690, p < 0.001, η² = 0.030), with extracurricular activities emerging as a significant covariate. Differences in psychological distress and burnout were particularly striking, with large effect sizes for DASS-21 (η² = 0.461) and School Burnout Inventory (η² = 0.402). Covariates such as mental health history, alcohol use, gender, sleep, and social media exposure also influenced these outcomes, highlighting the role of internal and external pressures. Clusters also varied substantially in their perception of the learning environment (η² = 0.418), influenced by health conditions, academic year, and sleep duration. Resilience dimensions showed some of the largest effects, particularly perseverance (η² = 0.512) and reflective/adaptive help-seeking (η² = 0.504), both linked to study habits and year of study. Coping strategies also differed markedly across clusters: problem-focused (η² = 0.368), avoidant (η² = 0.314), and emotion-focused coping (η² = 0.228). Covariates including extracurricular activity, mental health history, alcohol consumption, and gender significantly shaped coping responses, with avoidant coping especially elevated among those with prior mental health conditions or alcohol use. Collectively, these findings highlight the multifaceted psychological and behavioral distinctions between clusters.

**Table 2.** ANCOVA Results for Psychological Profiles with Clusters as Independent Variable and controlling for covariates.

| Dependent Variables | F-value | P-value | Partial $\eta^2$ | Significant Covariates |
| --- | --- | --- | --- | --- |
| Grade Point Average | 6.690 | P<0.001*** | 0.030 | Extracurricular Activities |
| Depression, Anxiety, and Stress Scale - 21 | 188.369 | P<0.001*** | 0.461 | Mental Health Diagnosis, Alcohol Consumption, Gender, Year, Average Sleep Time, Average social media Time |
| School Burnout Inventory | 147.894 | P<0.001*** | 0.402 | Year, Average Social Media Time |
| Dundee ready Educational Environment Measure | 158.160 | P<0.001*** | 0.418 | Chronic Diseases, Year, Average Sleep Time |
| Perseverance (ARS) | 230.767 | P<0.001*** | 0.512 | Year, Average Study Time |
| Reflective and Adaptive Help Seeking (ARS) | 223.374 | P<0.001*** | 0.504 | Average Study Time |
| Negative Affect and Emotional Response (ARS) | 208.918 | P<0.001*** | 0.487 | Mental Health Diagnosis, Average Sleep Time, Average social media Time |
| Problem Focused (Brief - COPE) | 127.952 | P<0.001*** | 0.368 | Extracurricular Activities |
| Emotion Focused (Brief - COPE) | 64.487 | P<0.001*** | 0.228 | None |
| Avoidant (Brief - COPE) | 100.849 | P<0.001*** | 0.314 | Mental Health Diagnosis, Alcohol Consumption, Gender |

Adjusted means and post-hoc comparisons (Figure 1D; Table 3) showed that Cluster D had the highest GPA and Cluster C the lowest, with the largest gap observed between these two groups (p < 0.001). Clusters A and B both have relatively high GPA levels, but with distinct psychological profiles. A notable pattern is in burnout and stress, where both A and C scored highly, with C showing the greatest distress. However, their coping patterns diverged, with A ranked among the strongest in resilience and adaptive coping, whereas Cluster C was lowest across all groups. Another notable contrast is that although A and B were the most resilient, B reported the lowest stress overall. Cluster D was consistently moderate across measures. Perceptions of the learning environment were most positive in B and weakest in C. These results underscore that although Clusters A and C both face high stress, their ability to adapt markedly differs, pointing to different support needs.

**Table 3.** Pairwise Comparison Matrix of Various Feature Characteristics of the Clusters.

| Dependent Variables<sup>a</sup> | Cluster A - Cluster B | Cluster A - Cluster C | Cluster A - Cluster D | Cluster B - Cluster C | Cluster B - Cluster D | Cluster C - Cluster D |
| --- | --- | --- | --- | --- | --- | --- |
| GPA | 0.43 | 0.028* | 0.08 | 0.224 | 0.045* | $P < 0.001^{***}$ |
| DASS21 | $P < 0.001^{***}$ | 0.01* | $P < 0.001^{***}$ | $P < 0.001^{***}$ | $P < 0.001^{***}$ | $P < 0.001^{***}$ |
| SBI | $P < 0.001^{***}$ | $P < 0.001^{***}$ | $P < 0.001^{***}$ | $P < 0.001^{***}$ | $P < 0.001^{***}$ | $P < 0.001^{***}$ |
| Problem Focused (Brief-COPE) | 0.606 | $P < 0.001^{***}$ | $P < 0.001^{***}$ | $P < 0.001^{***}$ | $P < 0.001^{***}$ | $P < 0.001^{***}$ |
| Emotion Focused (Brief-COPE) | $P < 0.001^{***}$ | $P < 0.001^{***}$ | $P < 0.001^{***}$ | 0.204 | $P < 0.001^{***}$ | 0.039* |
| Avoidant (Brief-COPE) | $P < 0.001^{***}$ | 0.016* | $P < 0.001^{***}$ | $P < 0.001^{***}$ | 0.016* | $P < 0.001^{***}$ |
| DREEM | $P < 0.001^{***}$ | $P < 0.001^{***}$ | $P < 0.001^{***}$ | $P < 0.001^{***}$ | $P < 0.001^{***}$ | $P < 0.001^{***}$ |
| Perseverance (ARS) | $P < 0.001^{***}$ | $P < 0.001^{***}$ | $P < 0.001^{***}$ | $P < 0.001^{***}$ | $P < 0.001^{***}$ | $P < 0.001^{***}$ |
| Reflective and Adaptive Help Seeking (ARS) | $P < 0.001^{***}$ | $P < 0.001^{***}$ | $P < 0.001^{***}$ | $P < 0.001^{***}$ | $P < 0.001^{***}$ | $P < 0.001^{***}$ |
| Negative Affect and Emotional Response (ARS) | $P < 0.001^{***}$ | $P < 0.001^{***}$ | $P < 0.001^{***}$ | $P < 0.001^{***}$ | $P < 0.001^{***}$ | $P < 0.001^{***}$ |

<sup>a</sup> P-value for pairwise comparison was calculated using Holm's correction

### Qualitative Validations

The FGDs confirmed the distinctiveness of the four clusters while providing depth to their psychological profiles (Table 4). Cluster A students described stress spikes during assessments and remedials, often amplified by stigma (“*‘You remediated?’… I was disappointed, but it became a lesson*”). Yet this stigma frequently triggered renewed effort, supported by peer study circles, family encouragement, and faith-based coping. These explain how this cluster maintains academic performance despite the high stress levels. Cluster B students reflected a thriving profile, with stress reframed as part of the learning curve (“*Without stressors, we get lazy… the right amount makes us enthusiastic*”). They emphasized personal growth, involvement in student organizations, and a generally supportive environment. While both A and B relied on peer study circles, A used them to manage stress, whereas B used them to reinforce their thriving performance.

**Table 4.** Psychological Characteristics of each Cluster Evaluated from Quantitative and Qualitative Analysis.

| Clusters | Quantitative Characteristics | Qualitative Themes | Representative Quotes |
| --- | --- | --- | --- |
| **Cluster A** | High burnout and psychological stress<br>High resilience<br>Strong perception of their learning environment<br>High adaptive and maladaptive coping scores. | High stress during initial exams (1 and 2)<br>Various adaptive, help-seeking, and faith-based coping (3, 4, and 5)<br>Environment generally supportive (5 and 6) | "I cried at first... 'you remediated?'... disappointed... but it became a lesson."<br>"My mom said, 'Try to calm yourself... I use breathing... count and slow your breathing.'"<br>"Eventually, I went for treatment at the university's hospital... the doctor, also a hypnotherapist, suggested either psychiatry or hypnotherapy. I chose hypnotherapy... and after the sessions, I was functional again in daily life."<br>"I rely on myself... I rarely confide in friends—I confide directly in God. When I pray, it's never formal... 'God, today <redacted> has this... what should I do?' That way, my heart feels calmer... when I read scripture, I feel more at peace."<br>"If the tutor explains well, it's really good... but if they stay silent, it becomes a barrier... we need tutors who are willing to explain."<br>"It helps, actually... my academic advisor encouraged me—'It's okay, keep up the spirit'... that kind of support really helps." |
| **Cluster B** | Low burnout and psychological stress<br>High resilience<br>Strong perception of their learning environment<br>High adaptive coping and low maladaptive coping scores. | Thriving in medical schools (1 and 2)<br>Coping through reframing and acceptance of stress (3, 4, 5, and 6)<br>Peers and Student Organizations are Important (2 and 7)<br>The environment is seen as supportive (7 and 8) | "Actually, I don't really get stressed... what matters is that I've studied properly."<br>"Through joining organizations, I felt I grew... I discovered a new version of myself through medical school."<br>"If I remedial even though I mastered the cases, that's beyond me... so I just accept it."<br>"As long as I've tried, that's enough... what satisfies me is the learning process, not just the exam result."<br>"Without stressors, we get lazy... too much causes demotivation. But the right amount makes us enthusiastic."<br>"The Filipi–Sinai split (Context: during exams, students were stratified by their current GPAs as reflected by the room Filipi or Sinai) makes stratification obvious, but it also motivates us to prove ourselves."<br>"Joining the student organization really helped—my grades improved, and that's when I first made the dean's list."<br>"My academic advisor is very supportive... even just during signatures, they gave encouragement." |
| **Cluster C** | High burnout and psychological stress<br>Low resilience<br>Weak perception of their learning environment<br>Low adaptive coping and high maladaptive coping scores. | Medical schools were not their first choice (1, 2, and 3)<br>Stress translates to self-blame (4 and 6)<br>Maladaptive coping and helplessness (4, 5, 6, and 12)<br>Students request higher | "Actually, medicine was never my goal... it was more my parents' choice. I just go through it, but it feels exhausting."<br>"From the start, I didn't want to be in medical school... if not for my study circle friends, I don't think I could endure it."<br>"I entered medicine just by following along... my parents told me to, so I did." |
|  |  | support from the environment (7, 8, and 9)<br>Peers are double-edged (2, 10, and 11) | But the path is really heavy."<br>4. "When I fail, I just cry, blame myself... then lose motivation to start again."<br>5. "I once skipped an OSCE because I was too anxious. It felt easier not to come than to face it."<br>6. "When I get sanctioned or fail, I just shut myself in my room... I reflect a little, but end up procrastinating and repeating mistakes."<br>7. "Sometimes lecturers just read the slides... It's no different from studying alone."<br>8. "Most academic advisors are passive... just sign forms, rarely truly helpful."<br>9. "I often learned more from my study circle... because lectures often didn't stick."<br>10. "I'm often compared with those with higher grades... it's very pressuring, I'm afraid of being left behind."<br>11. "I once heard rumors about me (from my circle of friends)... it really made me feel inferior."<br>12. "I was confused where to seek help... if it were available at the university, it would be much easier." |
| **Cluster D** | Low burnout and psychological stress,<br>Moderate resilience,<br>Moderate perception of their learning environment,<br>Low adaptive and maladaptive coping scores. | Stress is manageable and not overwhelming (1 and 2)<br>Practical stress reduction techniques (3, 4, and 5)<br>Support is adequate but can be improved (6 and 7)<br>Issues about fairness (8 and 9) | 1. "When I'm tired, I just sleep, play games, or listen to music... it never reaches stress."<br>2. "At first, exams made me nervous... but after trying it, I learned the trick. Since then, I don't feel stressed, just adapt."<br>3. "Before exams, I cried daily... but later I learned to manage emotions with breathing techniques and small things that made me happy."<br>4. "At first I cried when I was late submitting a practical... but then I learned to manage tasks better and set aside rest time."<br>5. "I cope by exercising and hanging out with friends... it actually makes studying easier."<br>6. "Some lecturers really motivated me to learn more... but others made me lose interest; the material felt less motivating."<br>7. "The skills lab helps... but sometimes mannequins don't work properly."<br>8. "Examiners differ... some pass easily, some don't, even if we studied the same."<br>9. "One lecturer teaches us one way, another makes the exam... the answers end up different." |

Cluster C students conveyed heightened vulnerability, noting that medicine was often not their choice (“*Actually, medicine was never my goal… it was my parents’ choice*”). Passive teaching and a lack of autonomy appeared to erode resilience, leading to self-blame and maladaptive coping (“*When I fail, I just cry, blame myself… then lose motivation*”). They requested clearer communication, more engaging pedagogy, and accessible support services, confirming their need for intensive interventions. Cluster D students were high performers with low stress, managed by simple routines (“*When I’m tired, I just sleep, play games, or listen to music… it never reaches stress*”). Unlike other clusters, they did not seek intensive support but highlighted fairness and transparency in assessments as central to their engagement. Taken together, the qualitative validation strongly aligned with the quantitative results, showing that each cluster differed not only in psychological measures but also in lived experiences, coping, learning environment, and support needs. These findings demonstrate that clustering can reveal distinct subgroups whose needs differ markedly, supporting the paradigm of precision student support.

## Discussion

Our findings highlight the utility of clustering in identifying distinct psychological profiles among medical students, reinforcing the need for tailored interventions rather than a one-size-fits-all approach. The clustering analysis revealed four psychologically and academically distinct student groups, with Clusters A and C characterized by high burnout and psychological stress, and Clusters B and D showing lower distress but different resilience and coping patterns. The differences observed across clusters persisted even after adjusting for significant covariates, including mental health diagnoses, sleep habits, and extracurricular participation. This suggests these clusters reflect inherent psychological differences rather than spurious or artefactual findings.

Targeted interventions on students were not a new finding. Several studies have shown that intervention effectiveness varies depending on student characteristics. ^35,36^ For example, a study by Sukhwant et al. identified that academic support interventions benefit low-achieving students better than high-achieving students.^37^ Supporting this, the MYRIAD cluster randomized controlled trial by Kuyken et al. explored the effectiveness of universal school-based mindfulness training (SBMT) and found no evidence that SBMT was superior to standard teaching approaches. ^38,39^ These findings highlight that targeted, broad, universal interventions may be less effective than targeted programs. Our study identified key characteristics of student clusters and several potential treatment targets for student clusters, aligning with their different characteristics.

One of our key findings is Cluster C, which represents the most at-risk student population. This cluster demonstrates high psychological distress, low resilience, and poor academic performance. Previous research has shown that students experiencing both high distress and low resilience are more likely to struggle academically, develop mental health issues, or disengage with the learning environment. ^40–45^ Given the vulnerabilities, these students might represent populations where multiple approaches may be necessary, including academic support, psychological support, stress reduction training, and academic resilience training. The qualitative findings reinforced these interpretations. Many students in the cluster described entering medicine due to parental pressure rather than personal choice (“*Actually, medicine was never my goal… it was more my parents’ choice. I just went through it, but it feels exhausting*”), which amplified feelings of disengagement and helplessness. Their coping styles were often maladaptive, with failures leading to self-blame and withdrawal (“*When I fail, I just cry, blame myself… then lose motivation to start again*”). Perceptions of the learning environment were also weaker than in other clusters, with frustration at passive teaching styles (“*Sometimes lecturers just read the slides… it’s no different from studying alone*”) and a lack of accessible support. Although exposed to the same environment, these students requested intensive academic and psychological support.

Findings from Cluster C can be further explored using several theoretical frameworks. The Self-Determination Theory emphasizes the role of autonomy, competence, and relatedness in sustaining motivation. ^46,47^ Many Cluster C students entered medicine due to parental pressure, reflecting that their choice lacked autonomy, a key driver of intrinsic motivation. Without a sense of ownership, their motivation is easily undermined, with studies showing links between motivation and student performance and well-being. ^48^ The Job Demands–Resources framework provides an additional explanation. According to this framework, high demands can be buffered by adequate personal and contextual resources, preventing strain and burnout. ^49^ Cluster C students face the same heavy demands as their peers but also perceive themselves as having fewer resources available within themselves, their peers, and their learning environment. As a result, their demands remain unaddressed. Studies have shown that without these resources, the demands lead to heightened distress, maladaptive coping, ^50^ and ultimately poorer academic performance. Together, these frameworks illustrate how a lack of autonomy and insufficient resources interact to produce this cluster’s demotivation and poor academic performance.

Cluster C also represents students who requested specific interventions tailored to them, including academic and psychological support (Table 4). Studies have shown that interventions targeting at-risk students have greater benefits than universal programs. ^35,36,51^ Furthermore, several interventions showed multiple benefits within a single program. For example, burnout mitigations were shown to improve students’ learning environment, stress level, and enhance academic performance, while peer support reduces students’ burnout and psychological stress and increases resilience.^40,44,52–57^ Both quantitative and qualitative findings indicate that Cluster C requires immediate and targeted intervention. In addition to individualized psychological counseling and resilience training, systemic improvements such as engaging pedagogy, clearer communication, and proactive academic advising may help reduce the risk of disengagement. This cluster underscores the need for institutions to go beyond generic wellness programs and prioritize tailored interventions for students facing the combined burden of psychological distress, low resilience, and weak environmental support.

Cluster A represents another at-risk population, characterized by students experiencing high burnout and psychological stress, yet demonstrating strong resilience and adaptive coping skills. Despite these, the students remain susceptible to burnout, which may negatively impact their academic performance over time. Academically, Cluster A students perform better than those in Cluster C, suggesting that interventions should focus on stress management before it affects their academic performance. Strategies such as stress reduction workshops and targeted psychotherapy could help prevent the long-term adverse effects of chronic stress exposure on academic performance. ^45,58,59^ Our qualitative analysis reinforced the quantitative results as students described acute stress spikes around high-stakes assessments and remedials, often amplified by stigma (“*‘You remediated?’… I was disappointed, but it became a lesson*”). However, despite the high stress conditions, they also showed deliberate adaptation, relying on peer study circles, family, and faith-based coping (“*I rely on myself… I rarely confide in friends—I confide directly in God. When I pray, it’s never formal… my heart feels calmer*”). These narratives show how Cluster A students develop resilience through external support and internal reframing, which can be explained using the Job Demands-Resources model. ^49^ Cluster A students face significant stressors similar to their peers in Cluster C. However, they actively draw upon multiple resources from peer networks, family encouragement, and faith-based coping, which buffer the impact of these demands. Studies consistently show that such resources can mitigate the effects of stress and burnout, ^50,60,61^ but this buffering reflects a dynamic and sometimes fragile balance between demands and resources. Longitudinal evidence indicates that resources only weakly affect burnout ^61^ and only buffer two (exhaustion and cynicism) out of the three burnout components. ^60^ Therefore, we argued that even well-mobilized resources eventually erode when demands remain high, leading to renewed strain and disengagement. Over time, institutional scaffolds are needed to replenish resources and sustain coping capacity, as tipping this balance could compromise resilience and academic performance.

In contrast, Clusters B and D represent high-achieving students with low levels of burnout but distinct resilience and coping levels. Qualitative findings reinforced this as Cluster B students consistently reframe stress as part of their growth (“*As long as I’ve tried, that’s enough… what satisfies me is the learning process, not just the exam result*”) and highlighting the role of student organizations in shaping their identity (“*Through joining organizations, I discovered a new version of myself in medical school*”). These accounts support the interpretation of Cluster B as a thriving group in medical schools. These findings from Cluster B can be explored through several frameworks, including the Self-Determination Theory ^46,47^ and the concept of Communities of Practice. ^62^ Students in this cluster experienced high levels of autonomy in their learning, a strong sense of competence, which appeared to originate from their inner confidence and deep feelings of relatedness through peer networks and student organizations. (Table 4) A study identified that satisfying these three basic needs promoted students to thrive, increasing their resilience and psychological well-being. ^63^ This might explain their ability to reframe stress as an opportunity, reflecting their higher resilience and engagement. These students also preferred student organizations and peer study circles, which align with the Communities of Practice concepts. ^62^ Studies have shown that within these learning communities, they can advance academically and also develop increasing social capital, ^62,64,65^ allowing them to further thrive in medical education. These also suggest that Cluster B students may be suited to serve as near-peer mentors. Several studies have linked peer mentoring to improvements in academic performance, empathy, and burnout reduction. ^53,57,66^ We argue that providing opportunities for them to cultivate leadership and mentoring roles may benefit both the mentor and the mentee.

Despite their high GPA, Cluster D students did not show statistically significant differences from Cluster B in pairwise comparisons of academic performance. Qualitative findings identified that these students adapted quickly and can manage stress with simple routines (“*When I’m tired, I just sleep, play games, or listen to music… it never reaches stress*”). These clusters did not request intensive interventions but emphasized fairness and transparency in assessments (“*Examiners differ… some pass easily, some don’t, even if we studied the same*”). This suggests that their concerns are more about expectations of fairness. Viewed through the lens of Organizational Justice,^67^ these findings suggest that Cluster D students, although psychologically stable, are sensitive to perceived unfairness in evaluation. Because they experience less stress from academic workload, simple coping strategies suffice. However, their engagement is undermined when fairness is questioned, leading to potential dissatisfaction despite strong performance. Studies have shown that perception of organizational justice significantly influences students’ attitudes and overall educational experience, ultimately determining their motivation and academic engagement. ^67–70^ This dynamic points to issues in assessment practices and highlights the need for reforms such as transparent criteria, consistent feedback, and examiner calibration. Complementary programs, including wellness initiatives, resilience training, and incremental improvements to assessment practices, may also support this group’s long-term well-being and engagement. ^16,71^ Although such approaches do not directly address fairness concerns, they can augment institutional reforms by strengthening coping capacity and sustaining resilience. Nevertheless, further research is needed to determine the most effective interventions for maintaining engagement and academic performance among these students.

Aside from psychological factors, the differences in academic performance between clusters can be interpreted through several educational theories. The sociocultural learning theory by Vygotsky emphasizes that learning occurs primarily through social and cultural interactions. ^72,73^ This theory is reflected in Cluster B, where students thrived through peer study circles and student organizational involvement, highlighting how social contexts support learning. In contrast, sociocultural learning was less active in Cluster C. Students in this cluster not only reported weaker peer support but also described peer learning as “double-edged,” which potentially undermines rather than enhances their opportunities for engagement. The concept of the zone of proximal development further explains these patterns. ^73,74^ Vygotsky proposed that learning is most effective when tasks lie just beyond a learner’s current ability but can be achieved with support from a more knowledgeable person. We argue that this process also requires students to believe in their capacity to succeed as per the self-efficacy theory by Bandura. ^75^ The theory explains that self-efficacy determines whether students will attempt and persist with challenging tasks. ^46,75^ In our findings, Cluster A students demonstrated strong coping and drew upon social and familial resources, while Cluster B students leveraged extensive networks and study groups, allowing both clusters to operate effectively within their zone of proximal development and enhancing their self-efficacy. By contrast, Cluster C students lacked such scaffolding and reported self-blame and low self-belief in the face of failure, leaving them unable to mobilize the support necessary to progress academically. Viewed through these theories, precision student support potentially enables students to develop the capacity to cultivate resources, whether scaffolding, confidence, or social interactions needed to thrive in medical schools.

This study has several limitations that should be considered. First, the study was conducted at a single institution, which may limit the generalizability of the results to other schools with different curricula, cultural contexts, or support systems. Multi-center studies are needed to determine whether the clusters are consistent across various settings. Second, the cross-sectional design prevents us from evaluating the dynamics of how students transition between clusters over time. Longitudinal follow-up could provide essential insights into the stability of cluster membership and the effectiveness of targeted interventions. Third, other clustering methods may yield slightly different groupings as clustering results are inherently sensitive to data structure and parameter choice. Fourth, the qualitative validation provided depth and triangulation of the quantitative clusters, but the sample was small, and therefore, the qualitative findings should be considered illustrative rather than representative. Finally, while this study highlights the potential of precision student support, implementation studies are still required to evaluate cluster-specific interventions’ feasibility and long-term effectiveness in real educational contexts.

Our findings highlighted the benefit of clustering analysis in distinguishing student populations that require tailored interventions. We identified four clusters, each with its own psychological features and needs. Cluster C students require immediate psychological and academic support, while Cluster A students may benefit from stress management and burnout prevention to sustain their resilience. Cluster B students could serve as peer mentors, leveraging their strengths to support others, while Cluster D students need interventions that promote long-term well-being. Our findings contribute to the emerging paradigm of precision education. Just as precision medicine tailors therapies to biological subtypes, precision student support adapts interventions to psychological and contextual subgroups. In contrast, traditional wellness programs applied universally to entire cohorts may dilute effectiveness and fail to engage students with divergent needs. For example, our qualitative findings revealed that while Cluster B students thrive and could serve as mentors, Cluster C students, who often entered medicine under parental pressure and expressed disengagement, require far more intensive and structured support. Recognizing such heterogeneity underscores the limitations of generic wellness initiatives and highlights the value of a precision-based framework in global debates on medical student mental health and educational sustainability. By adopting a data-driven approach to student support, institutions can better align resources with each cluster’s needs. Such targeted interventions can improve student well-being, sustain academic performance, and foster resilience in future healthcare professionals. Across these educational and psychological theories, the common idea is that student support must cultivate autonomy, resources, fairness, and scaffolding, aligning interventions with specific cluster needs. Future research should evaluate the long-term effectiveness of cluster-specific interventions and explore whether students transition between clusters over time. This could inform early detection and preventive strategies to mitigate stress, enhance resilience, and ensure sustainability in medical education.

## Author Contributions

Ardo Sanjaya contributed to conceptualization, methodology, data collection, formal analysis, interpretation, writing – original draft, writing – review and editing, supervision, and final approval. Christian Edwin contributed to conceptualization, data collection, formal analysis, interpretation, writing – original draft, and final approval. Cindra Paskaria contributed to data collection, formal analysis, interpretation, writing – review and editing, and final approval. Nathanael Andry Mianto contributed to data collection, interpretation, writing – review and editing, and final approval. Renata Alviani contributed to data collection, writing – original draft, and final approval. Kevin Gunawan contributed to data collection, writing – original draft, and final approval.

## Data availability statement

The datasets generated during the study are available from the corresponding author upon reasonable request and are subject to ethical considerations.

